# Joint Investigation of Two-Month Post-Diagnosis IgG Antibody Levels and Psychological Measures for Assessing Longer Term Multi-Faceted Recovery among COVID-19 Cases in Northern Cyprus

**DOI:** 10.1101/2020.08.27.20178160

**Authors:** Burc Barin, Banu Elcin Yoldascan, Fatma Savaskan, Goncagul Ozbalikci, Tugce Karaderi, Hüseyin Çakal

**Author notes:** co-senior authors (Tugce Karaderi, Hüseyin Çakal).

## Abstract

Following the outbreak of COVID-19, multidisciplinary research focusing on the long-term effects of the COVID-19 infection and the ‘complete longer term recovery’ is still scarce. With regards to long-term consequences, biomarkers of physiological effects as well as the psychological experiences are of significant importance for comprehensively understanding the whole COVID-19 recovery period. The present research surveys the IgG antibody titers and the impact of COVID-19 as a traumatic experience in the aftermath of the active infection period, around two months after diagnosis, in a subset of COVID-19 patients from the first wave of the outbreak in Northern Cyprus. Associations of antibody titers and psychological survey measures with baseline characteristics and disease severity were explored, and correlations among various measures were evaluated. Of the 47 serology tests conducted for presence of IgG antibodies, 39 (83%) were positive. We identified trends demonstrating individuals experiencing severe or critical COVID-19 disease and/or those with comorbidities are more heavily impacted both physiologically and mentally, with higher IgG titers and negative psychological experience compared to those with milder disease and without comorbidities. We also observed that more than half of the COVID-19 cases had negative psychological experiences, being subjected to discrimination and verbal harassment/insult, by family/friends. In summary, as the first study co-evaluating immune response together with mental status, our findings suggest that further multidisciplinary research in larger sample populations as well as community intervention plans are needed to holistically address the physiological and psychological effects of COVID-19 among the cases in the long-term.

## Section I. Introduction

Coronavirus disease of 2019 (COVID-19), resulting from SARS-CoV-2 infection, was declared a pandemic by the World Health Organization on 11 March 2020. As of 29 July 2020, more than 16,000,000 COVID-19 cases were identified, and more than 650,000 deaths were reported due to the disease (1). Although the scientific community has responded rapidly to detect the transmission mechanisms and develop vaccines, multidisciplinary research focusing on the long-term effects of the COVID-19 infection is still scarce, and not much is known on how the human body responds to COVID-19 infection, both biologically and psychologically during the ‘longer term recovery’ period after discharge from the hospital/isolation. With regards to long-term effects, biomarkers of physiological effects as well as the psychological experiences are of significant importance for a comprehensive understanding of the COVID-19 recovery period (2). COVID-19 as a life threatening infection can act as an acute stressor (3) and stress can have a down-regulatory effect on the immune system (4). The present research surveys the IgG antibody titers and the impact of COVID-19 as a traumatic experience both during and in the aftermath of the active infection period.

There is insufficient information on the immune response to COVID-19 (e.g. prevalence of different antibodies against the infection over time and development of long-term immunity). It is essential to better understand the timeline of immune response including the appearance of immunoglobulin M (IgM) and immunoglobulin G (IgG) antibodies, their lifespan and whether they are protective, at least partially, against a second infection. Preliminary research shows that detectable IgG antibodies generally start appearing after the first week after symptom onset, reach a peak around two to three weeks, and stay at detectable blood levels at least for a duration of 2-3 months even in milder cases, similar to previous observations in other SARS infections (5-7). Moreover, the psychological effects of having the infection are also complex. The risk of having severe COVID-19, the overall disease burden, along with many unknowns about its short- and long-term effects, increase the stigma attached to the infection and the related anxiety among the public, and make COVID-19 cases vulnerable to post-traumatic stress as well as targets for harassment and discrimination (8). It is presumed that the period of complete physiological and psychological recovery from the infection depends on disease severity and other physiological and socioeconomic factors. However, given all the elaborate aspects of COVID-19 yet to be investigated and understood, the long-term multi-faceted recovery period is still far from being deciphered.

From a psychological point of view, initial findings suggest that both the disease itself and the negative consequences of the lockdown imposed by governments to curb the spread of the disease could result in negative coping behaviour which includes but is not limited to panic, anxiety, stigmatization, and post-traumatic stress disorder (PTSD) (3). As scarce research shows, these reactions can also be influenced by contextual factors such as a history of war, famine, and the size of the population. More specifically, while smaller nations might appear to have the upper hand in rapid enforcement of measures, contextual factors such as the increased connectivity of the individuals in smaller societies, or negative collective experiences of war and famine in the past might increase the prevalence of negative coping behaviours and stigma induced depression (9).

A particular case in point is Northern Cyprus, governed by a state that remains internationally unrecognized, and hence, not included in the global epidemiological COVID-19 statistics. In the first wave of the COVID-19 outbreak in Northern Cyprus, 108 cases were diagnosed between 10 March and 16 April 2020. The authorities responded promptly and lockdown was imposed on March 11 (9) effectively halting education and government offices, and all other services except those considered essential. In addition to the global scare about the pandemic, the small community setting in Northern Cyprus (an estimated total population of around 400,000) with a history of war and trauma (10) further caused an intensified disease stigma. On the one hand, panic engulfed the small nation and there was widespread stigma toward those who tested positive or considered high-risk for transmitting the disease, i.e. Turkish Cypriots living abroad, who were brought home and quarantined (11). On the other hand, videos of individuals under duress as a result of being quarantined were widely circulated in the social media and the public reacted negatively (12). Similarly, those who were tested positive recounted psychological trauma as their names made public and have been targeted (13). Therefore, there is sufficient grounds to assume that in addition to the physiological impact of the disease, those who tested positive for the COVID-19 have also experienced psychological distress during and after the active infection period. In fact, in an earlier study conducted in Wuhan (China), the prevalence of significant post-traumatic stress symptoms associated with COVID-19 was estimated as 96.2% among clinically stable COVID-19 cases at discharge from quarantine (14). Taken together, these observations suggest that assessing the biological markers of physiological effects vis-à-vis negative psychological experiences of the COVID-19 cases is important for holistic management of COVID-19 patients from diagnosis to potentially complete physiological and psychological recovery.

The present research surveys the immune response (IgG antibody titers) and negative psychological experiences among the COVID-19 cases in the extended recovery period in the small society setting of Northern Cyprus.

## Section II. Sample Population and Methodology

### Participants and Study Design

We performed a joint investigation of the immune response and mental status of the COVID-19 cases at an average time of two months after diagnosis. These two main outcomes of interest comprise the longer term recovery of the cases. Of the 108 cases diagnosed, 32 were tourists on the island: two died with the disease, and the remaining 30 individuals returned to their country after discharge. Of the remaining 76 individuals residing in Northern Cyprus, two died with the disease. A total of 74 individuals were invited to participate in the post-discharge assessment of antibody development and psychological impact. For the psychological evaluation, eight individuals under the age of 18 as well as three individuals who did not speak Turkish/English fluently were excluded from the study. Hence, a total of sixty-three individuals were eligible to participate in the psychological evaluation.

All subjects were informed about both components of the study, provided informed consent acknowledging voluntary participation, option to withdraw from study at any time, and the confidentiality of their antibody results and their responses to the survey.

### Eligibility Criteria

*General Inclusion Criteria:* Confirmed (i.e., with positive polymerase chain reaction test result) COVID-19 infection in Northern Cyprus between the dates of 10 March – 17 April and residence in northern Cyprus.

*Exclusion Criteria for Antibody Development Analysis:* Refusal to give informed consent, or contraindication to venipuncture.

*Exclusion Criteria for Psychological Survey:* Refusal to give informed consent, inability to understand/speak Turkish or English fluently, or being under the age of 18.

### Blood Collection and Transfer

Blood samples were taken by trained nurses during home visits. Venipuncture was used to collect blood. 10ml complete gel barrier formation tubes were used for blood collection (See Supplementary Text for the details).

### Serology Testing

The SARS-CoV-2 IgG assay is a chemiluminescent microparticle immunoassay (CMIA) intended for both the quantitative and qualitative detection of IgG antibodies to the nucleocapsid protein of SARS-CoV-2 in human blood serum and plasma. Assay specifications indicate that the SARS-CoV-2 IgG assay is intended for use as an aid in identifying individuals with an adaptive immune response to SARS-CoV-2, indicating recent or prior infection. This assay is only for use under the Food and Drug Administration’s Emergency Use Authorization. Per the assay’s recommended definition, we defined positive IgG response in the study as a titer level ≥ 1.4 index signal/cutoff (s/co) (15). Assays were run on Abbott’s ARCHITECTPlus i2000_SR_ System.

The reported positive predictive agreement (PPA) for the assay at ≥14 days post-symptom onset was 100.0% (95% confidence-interval (CI): 95.9%-100%) while the negative predictive agreement (NPA) was 99.6% (95% CI: 99.1%-99.9%). Performance characteristics of the assay were independently evaluated in a study conducted in Boise, Idaho, where specificity and sensitivity were reported as 99.90% and 100% (starting at day 17 after symptom onset), respectively (16).

### Psychological Measures

Whenever possible, we adapted and used already tested and validated measures to assess the negative psychological experiences of the cases. More specifically, we assessed experiencing COVID-19 as a life changing trauma, negative emotions, perceived importance of preventive measures, awareness and habits, initial reaction to diagnosis, evaluation of general health, stigma, perceived discrimination, post-traumatic anxiety, and evolving perspectives after discharge. Ordinal response scales with five levels were used for each question. Higher values indicated stronger experience of COVID-19 as a life changing trauma, perceived higher importance of preventive measures, stronger initial reaction to diagnosis, more positive evaluation of general health, more perceived discrimination, higher post-traumatic anxiety, and stronger anticipation of future COVID-19 related anxiety.

We assessed the internal reliability of our multi-item measures via Chronbach’s alpha (α>0.70; See Supplementary Material - Psychological Survey for the full Questionnaire). Experiencing COVID-19 as a life-changing traumatic was measured with three items (α=0.84) adapted from (17). Negative emotions during the recovery were assessed by four items (α=0.79). Perceived discrimination on the basis of being COVID-19 positive was measured by six items (α=0.90) adapted from (18). We also measured anxiety related to anticipated stigma in the future as a result of COVID-19 diagnosis with two items (r=0.82, p<0.001). We measured subjective evaluation of health before the diagnosis and after the discharge with a single item each. Willingness to help others by sharing information was measured by a single item and perceived importance of protective measures by 4 items (α=0.96).

### Statistical Analysis

Analysis of quantitative IgG titers was conducted via non-parametric tests: Wilcoxon rank-sum test (for factors with two levels) and Kruskal-Wallis test (for factors with three or more levels). We computed descriptive statistics for the socio-demographics factors and summary measures (mean score (M) with standard deviation (SD)) for psychological processes, and conducted Pearson correlation tests to explore whether the selected psychological processes were associated with each other. All multi-item measures and single items use 5-point Likert scales (1 lowest, 5 highest) and have a mid-level at 2.5. Disease severity was defined as critical (requiring intensive care), severe (requiring oxygen therapy, but otherwise stable) and mild/moderate (all other cases). P-values less than 0.001 were displayed as “p<0.001”. Statistical significance was defined as p < 0.05. Multivariate analyses were not carried out due to small sample size. Analyses were performed using SAS version 9.4 (SAS, Cary, NC, USA).

## Section III. Results

### Baseline Characteristics

Of the 74 cases eligible for serology testing, 47 (64%; 60% women and 40% men) accepted the invite and provided blood for testing. Median [interquartile range (IQR)] time from initial COVID-19 diagnosis to blood draw for serology testing was 66 [63.5-73] days with min-max of 50-86 days. Of the 63 cases eligible for responding to the psychological survey, 41 (65%) responded to survey questions (**Table 1**).

**Table 1.** Baseline Characteristics and Disease Severity by Endpoint

|  |  | **Serology**<br>**(N=47)** | **Psychological**<br>**Survey**<br>**(N=41)<sup>1</sup>** |
| --- | --- | --- | --- |
| **Sex** | Women | 28 (60%) | 23 (56%) |
|  | Men | 19 (40%) | 18 (44%) |
| **Age** | 0-29 | 9 (19%) | 7 (17%) |
|  | 30-59 | 25 (53%) | 21 (51%) |
|  | 60+ | 13 (28%) | 13 (32%) |
| **Education Completed** | Elementary School | 11 (23%) | 10 (24%) |
|  | Middle/High School | 15 (32%) | 13 (32%) |
|  | University or Higher | 18 (38%) | 18 (44%) |
|  | Not Reported | 3 (6%) |  |
| **Any Symptom**<br>**Reported at the Time**<br>**of Diagnosis** | No | 10 (21%) | 8 (20%) |
|  | Yes | 37 (79%) | 33 (80%) |
| **Fever/History of**<br>**Fever Reported at the**<br>**Time of Diagnosis** | No | 25 (53%) | 23 (56%) |
|  | Yes | 22 (47%) | 18 (44%) |
| **Comorbidity<sup>2</sup>** | No | 32 (68%) | 28 (68%) |
|  | Yes | 15 (32%) | 13 (32%) |
| **Disease Severity<sup>3</sup>** | Mild/Moderate | 38 (81%) | 33 (80%) |
|  | Severe/Critical | 9 (19%) | 8 (20%) |

<sup>1</sup> Of the 47 individuals who provided blood samples for serology testing, four cases were not invited to respond to the survey (one <18 years old, and three not fluent in local language) and two declined to participate in the survey.
<sup>2</sup> Most frequently reported chronic diseases were hypertension (N=9) and diabetes (N=5, two with concurrent hypertension).
<sup>3</sup> Disease severity was defined as critical (requiring intensive care), severe (requiring oxygen therapy, but otherwise stable) and mild/moderate (all other cases including asymptomatic cases).

For the serology testing, 19% were <30 years of age, 53% were between 30-60 years old, and 28% were ≥60 years of age. At the time of COVID-19 diagnosis, 79% and 47% of the serology analysis participants reported ‘at least one symptom’ and ‘fever history’, respectively. Thirty-two percent had at least one comorbidity – most frequently hypertension (N=9) and diabetes (N=5, two with concurrent hypertension). COVID-19 disease severity was severe or critical for 9 (19%) cases and mild/moderate for the remaining 38 (81%). For the psychological survey, distributions of participant baseline and disease severity characteristics were similar to those of the blood serology analysis (**Table 1**). Detailed cross-tabulation of baseline characteristics and disease severity by age group is displayed in **Supplementary Table 1**.

### Serology

Of the 47 serology tests conducted for IgG antibody development, 39 (83%) were positive and 8 (17%) were negative. All of the negative results came from individuals who experienced mild/moderate disease. Overall median [IQR] titer level was 4.38 [2.05-5.88]. Median [IQR] titer level among positives and negatives were 4.95 [3.79-6.09] and 0.61 [0.16-0.72], respectively.

**Figure 1** and **Supplementary Table 2** display the distribution of IgG antibody titers by baseline characteristics and disease severity. The factor that had the most impact on IgG titer at a median follow-up of two months post-diagnosis was disease severity. Nine subjects who had severe/critical disease had median [IQR] IgG titer of 6.09 [5.88-6.24] versus 3.94 [1.70-5.52] reported for thirty-eight subjects with mild/moderate disease (Wilcoxon rank-sum test; p=0.001). Among the baseline factors, fever/history of fever reported at the time of diagnosis yielded median [IQR] IgG titer of 5.56 [4.11-6.20] versus 3.57 [1.47-5.13] reported for those without fever/history of fever (Wilcoxon rank-sum test; p=0.01). Having a comorbidity also produced higher median [IQR] IgG titers of 5.52 [4.31-6.09] versus 3.87 [1.25-5.56] in those without a comorbidity (Wilcoxon rank-sum test; p=0.03).

**Figure 1.**
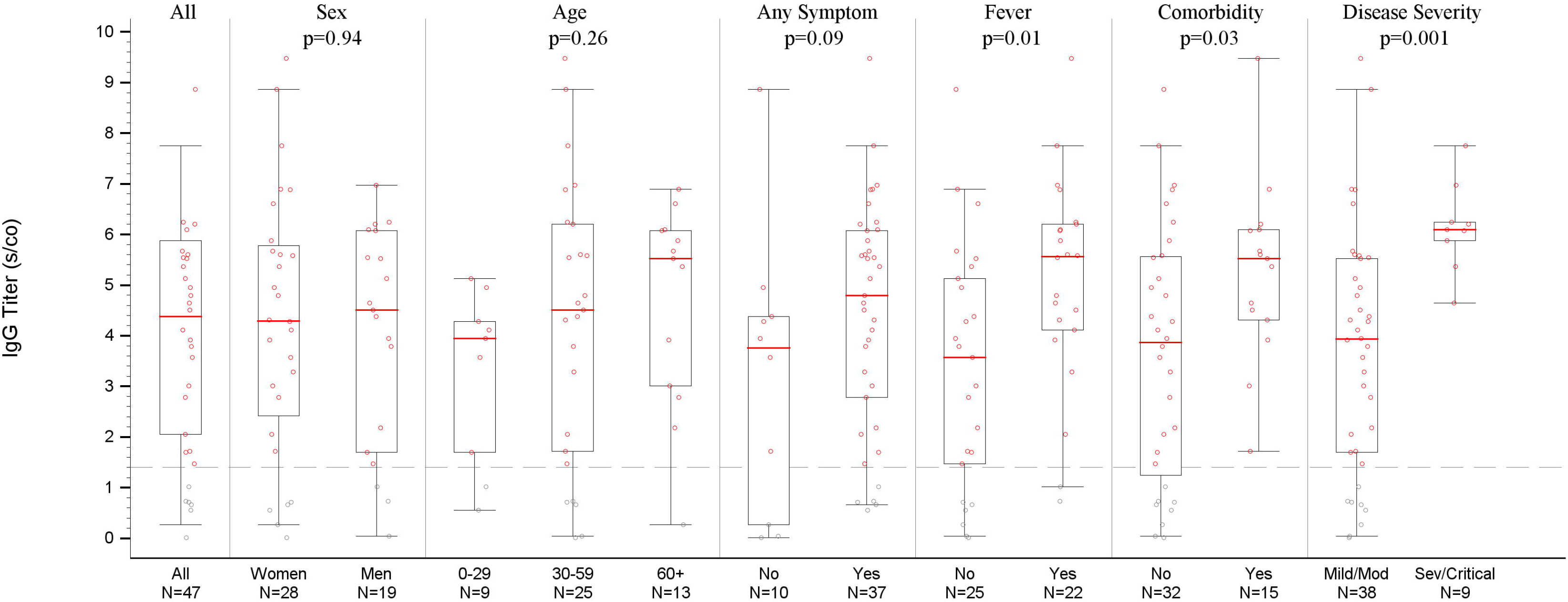
**Legend**. IgG Levels Overall, by Baseline Factors and Disease Severity. IgG levels were measured at a median [IQR] time of 66 [63.5-73] days from initial COVID- 19 diagnosis. See Supplementary Table 2 for detailed summary statistics. Mod=Moderate, Sev=Severe.

**Table 2.** Descriptive Statistics and Correlations Between the Measured Psychological Processes

| Process | M | SD | 1 | 2 | 3 | 4 | 5 | 6 | 7 | 8 |
| --- | --- | --- | --- | --- | --- | --- | --- | --- | --- | --- |
| 1. COVID-19 as Life-changing Trauma (CALCT) | 3.17 | 1.41 |  |  |  |  |  |  |  |  |
| 2. Negative Emotions | 2.61 | 1.25 | 0.54<sup>**</sup> |  |  |  |  |  |  |  |
| 3. Perceived Discrimination | 2.48 | 1.30 | 0.54<sup>**</sup> | 0.24<sup>ns</sup> |  |  |  |  |  |  |
| 4. Global Health before Diagnosis | 4.45 | 0.72 | -0.02<sup>ns</sup> | -0.07<sup>ns</sup> | 0.09<sup>ns</sup> |  |  |  |  |  |
| 5. Global Health after Diagnosis | 4.21 | 0.83 | -0.15<sup>ns</sup> | -0.20<sup>ns</sup> | -0.17<sup>ns</sup> | 0.42<sup>**</sup> |  |  |  |  |
| 6. Pro-social Tendencies | 4.39 | 0.97 | 0.25<sup>ns</sup> | 0.28<sup>ns</sup> | 0.18<sup>ns</sup> | -0.20<sup>ns</sup> | -0.16<sup>ns</sup> |  |  |  |
| 7. Perceived Importance of Protective Measures | 4.42 | 1.00 | 0.24<sup>ns</sup> | 0.09<sup>ns</sup> | 0.09<sup>ns</sup> | 0.14<sup>ns</sup> | -0.12<sup>ns</sup> | 0.41<sup>**</sup> |  |  |
| 8. Future Stigma Related Anxiety | 1.99 | 1.06 | 0.54<sup>**</sup> | 0.05<sup>ns</sup> | 0.80<sup>**</sup> | -0.07<sup>ns</sup> | -0.06<sup>ns</sup> | 0.13<sup>ns</sup> | -0.02<sup>ns</sup> |  |

M=Mean, SD=Standard Deviation
Note. Standardized coefficients are shown. Scales went from 1 (strongly disagree) to 5 (strongly agree). \*\* $p < 0.001$ ; ns: non-significant ( $p \geq 0.05$ ).

The distributions of IgG titers by cross-tabulation of baseline characteristics and disease severity are displayed in **Supplementary Table 3**. In the mild/moderate disease severity group, a significantly higher level of IgG titer was observed in individuals with comorbidities (median [IQR]: 5.02 [3.92-5.67]) compared to those without (median [IQR]: 3.43 [0.88-4.87]) (Wilcoxon rank-sum test; p=0.03).

### Negative Psychological Experiences

We report the descriptive statistics and the associations between negative psychological processes in **Table 2**.

Perception of COVID-19 diagnosis as a life changing traumatic event (CALCT) revealed a mean score of 3.17 [SD 1.41], which is above the mid-level. **Figure 2** displays the distribution of CALCT scores by baseline characteristics and disease severity. Similar to the IgG titer analysis, the factors that have shown trends for the most impact on CALCT scores at a median follow-up of two months post-diagnosis was disease severity, followed by presence of a comorbidity. Mean (SD) CALCT scores in mild/moderate and severe/critical disease groups were 3.01 (1.38) and 3.95 (1.42), respectively (Wilcoxon rank-sum test; p=0.10). For individuals with a comorbidity, mean (SD) CALCT score was 3.53 (1.48) as compared to 3.02 (1.39) in those without (Wilcoxon rank-sum test; p=0.30).

**Figure 2.**
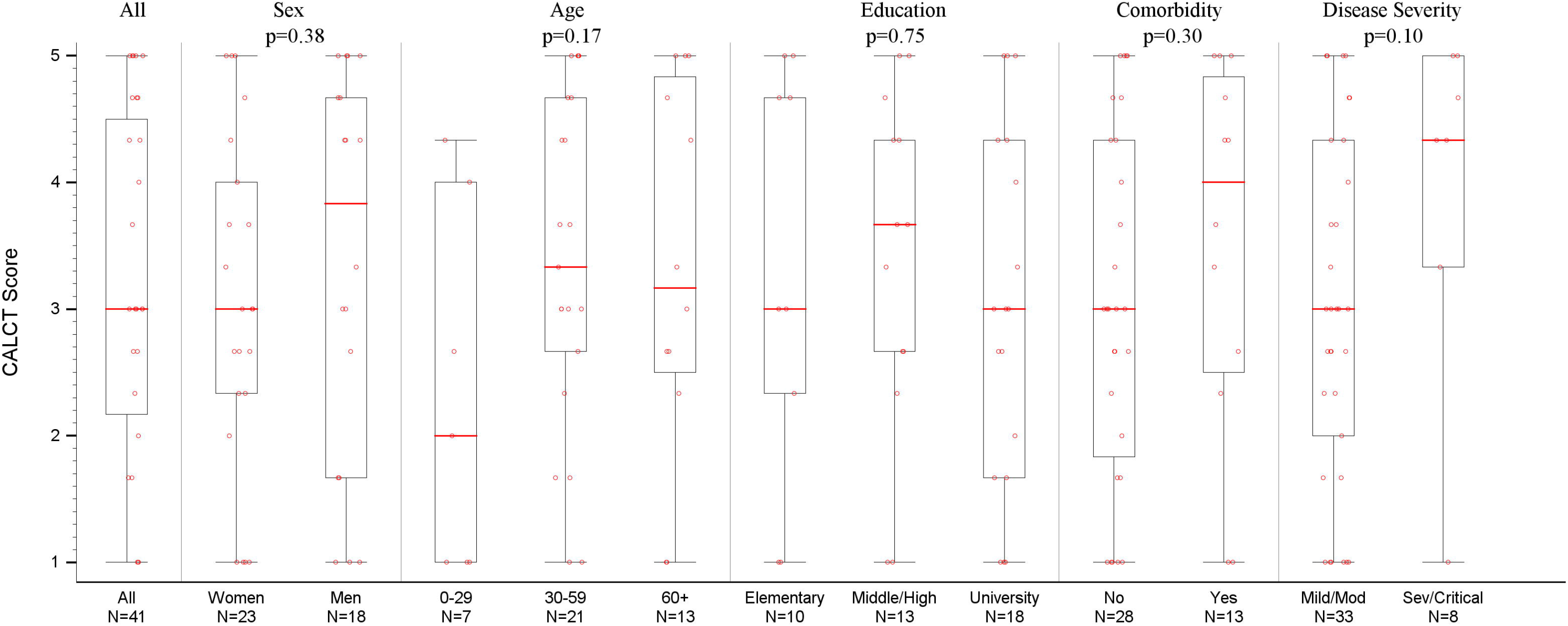
**Legend**. COVID-19 as Life-changing Trauma (CALCT) Scores Overall, by Baseline Factors and Disease Severity. Psychological measures were assessed around 2 months from initial COVID-19 diagnosis. See Supplementary Table 4 for distribution of responses to the three CALCT survey items. Mod=Moderate, Sev=Severe.

Among the individual questions measuring the negative psychological experiences, twenty-four (59%) respondents indicated a change in their perspective on life and their priorities due to the COVID-19 infection. All six (100%) responders to the question with severe/critical disease and 18 out of 33 (55%) responders with mild/moderate disease indicated a change in their perspective on life and their priorities due to the COVID-19 infection. 19 (46%) individuals indicated that they have become a more worried/anxious person because of the infection, and 20 (49%) perceived the infection period as a turning point in their lives (42% and 75% of the individuals with mild/moderate and severe/critical disease, respectively) (**Supplementary Table 4**).

The mean score for the negative emotions due to COVID-19 diagnosis was 2.61 (SD 1.25) and also above the mid-level of the scale (2.5). As for the individual emotions, felt as an initial reaction to COVID-19 diagnosis, worry ranked the first with 71% of respondents having felt it moderately, a lot or quite a lot, followed by helplessness (47%), fear of death (31%) and guilt due to not being sufficiently self-protected (19%). Fear of death and helplessness were both reported moderately or above by 27% and 50% of individuals in the mild/moderate and severe/critical disease severity groups, respectively (**Supplementary Table 5**).

Additional analyses of our psychological measures revealed that perceiving COVID-19 as a life changing trauma is strongly and positively associated with experiencing negative emotions (r=0.54, p<0.001); perceived discrimination (r=0.54, p<0.001); and future stigma related anxiety (r=0.54, p<0.001). Similarly, perceived importance of protective measures is again strongly and positively associated with pro-social tendencies (r=0.41, p<0.001). Last but not least, perceived discrimination at present is strongly and positively associated future stigma related anxiety (r=0.80, p <0.001) (**Table 2**).

## Section IV. Conclusions

We detected IgG antibodies in 39 (out of 47; 83%) of cases after a median of 66 days, which was a considerably longer follow-up period compared to the previous serological studies on IgG (on average up to ~30 days; 19-21). This observation confirms that IgG antibodies are still detectable in the blood in most COVID-19 cases around 2 months post-diagnosis. However, further studies are necessary to determine the neuralizing activity of these antibodies and whether they provide any immunity against a second infection. Moreover, severe/critical COVID-19 cases most of whom were older and/or with comorbidities had higher IgG titers, and also showed trends for the most impact mentally. Overall, we conclude that more specialized attention should be paid to this group for providing further monitoring and treatment post-discharge because of their higher healthcare needs related to comorbidities as well as the psychological impact in order to expedite the full recovery period after the infection.

Our analyses replicated the previous observations that disease severity is an important predictor of blood IgG levels (19-21), and confirmed that this observation holds true in the longer follow-up period we examined. Furthermore, among individuals with mild or moderate disease, we observed that those with comorbidities had significantly higher IgG levels (**Supplementary Table 3**). Similarly, Liu *et al*. observed that besides the severe COVID-19 cases who tended to have a more vigorous IgG response, a subset of the cases with mild disease had a robust IgG antibody response, and suggested that age and comorbidities may impact the timing and magnitude of the immune response (21). Fever reported at the time of diagnosis also hinted at a possible association with post-discharge IgG levels, but studies with larger case numbers are needed to evaluate these potential predictors of IgG levels with respect to potential confounders such as age, sex, different types of co-morbidities (e.g. autoimmune and endocrine-related diseases) and disease severity via multivariate models. All these factors with potential association to higher IgG titers are correlated with each other, and reflect increased disease burden during diagnosis and post-discharge (**Supplementary Table 1**). It is known that severity of COVID-19 is associated with a dysregulated immune response, and hence, further investigation of how dysregulated immune response is reflected in the long-term blood antibody levels may provide insights into the biological mechanism of the disease and support development of effective vaccines that are based on long-term immune response (22, 23).

In line with previous research, one in every two individuals with severe/critical disease felt fear of death and helplessness while one in every four individuals with mild/moderate disease felt these two emotions. Worry was the most commonly expressed emotion among the five emotions queried, with 71% of respondents having felt it moderately or more. Based on the responses to the psychological survey about two months after diagnosis, we infer that most cases have not yet recovered from the mental impact of the disease. Participants experienced COVID-19 as a life-changing trauma, experienced negative emotions, perceived themselves as discriminated against and experienced anxiety due to anticipated stigma in the future. In addition to replicating previous research on the negative psychological consequences of being tested positive for an infectious disease and that pandemics have a lasting negative impact on mental health among the general population (24-26), our findings also show that cases experienced anxiety as a result of anticipated stigma. This is a novel finding which reveals that pandemics like COVID-19 have long-term negative mental health effects. Future research could replicate and extend these findings via longitudinal designs.

Post-traumatic stress is an important part of this disease due to its overall severity, global impact and stigma attached to it. About half of the survey respondents reported being a more worried person due to the infection, and perceiving the infection as a turning point in their life. About one in four individuals also reported concern that their relationship with their workplace and family/friends will deteriorate due to infection. Hence, community resources for provision of psychological support to the COVID-19 cases post-discharge is very important to minimize the long-term impact of the disease and maintain mental health in these individuals. In Northern Cyprus, a number of organizations and universities have already taken action and set up psychological counseling hotlines, free for use by the public (9). These initiatives are very important and need to be expanded throughout the counties, regions and globally. However, more tailored intervention programs are needed especially for COVID-19 cases to combine mental check-ups with regular health check-ups at regular intervals. About one in ten individuals thought they could still transmit the disease. This provides another example of importance of using up-to-date medical info about the disease, and the person’s current status in providing tailored therapy to the person for getting over pre-conceived notions about fear of continuing disease in the individual.

Overall perception about the disease as a threat varied with disease severity (**Supplementary Table 12**). While more than half of the cases with mild/moderate disease deemed the infection was nothing to be afraid of, only one in four thought the same among the severe/critical cases. Therefore, a consistent public communication strategy is needed to ensure public perception of the disease will not change over time from a conscious alertness to the disease being ‘nothing to be afraid of due to sharing of experiences/perceptions by an estimated 80% of the cases in the mild/moderate severity group among the community.

The study is subject to a number of limitations. Although our study participation rates of 64% (serology) and 65% (psychology) among discharged COVID-19 cases are acceptable for an exploratory study such as this one. There may be some differences between individuals willing and unwilling to participate in the study, especially with respect to psychological endpoints. Actually, we observed lower participation rates in the study by cases from a rural region that was more severely impacted by the outbreak and had to go under a regional quarantine for an extended time. Disease stigma, continuing worry, suspicion and mistrust likely led to lower participation rates, and these factors are directly related to psychological endpoints studied here. To facilitate a more practical implementation in the field, it was not possible to use a consistent time point for evaluation of the outcomes of interest. Nevertheless, timing of blood draw and survey response showed limited variability around a two-month time point post-diagnosis, with median [IQR] and range time being 66 [63.5-73] and 50-86 days, respectively. There were possibly correlated responses for either or both endpoints as we allowed participation of multiple family or household members in the study. There were eight families that were represented in the study with 2-3 members each. Finally, compared to continuous IgG titer measures, categorical nature of the survey responses produced higher variability in calculated psychological process scores, and hence, lower statistical power in detecting any associations with baseline factors and disease severity.

In conclusion, this is the first study jointly evaluating post-discharge blood antibody levels and psychological status at a median time of two months after diagnosis. Severe/critical COVID-19 cases had higher blood IgG antibody levels as well as the highest long-term mental impact. Holistic and a more personalized approach is needed for post-discharge monitoring and treatment of COVID-19 cases, with a focus on older age, comorbidity status and disease severity.

## Data Availability

The datasets generated for this study are available upon request to the corresponding author.

## Acknowledgements

The authors would like to thank study participants as well as Savas Erdogan, Dr. Sebnem Benar, Safiye Ilgilier, Zeynep Unalan, Cigdem Adatas Sesen, Sumru Ozkan Ezer, Ayse Dogan for helping with the conduct of fieldwork and compilation of study data from psychological surveys, Sedef Kutlu and Serife Can from the microbiology laboratory running the immunoassays, Dr. Selin Özcem, Dr. Zafer Erdoğmuş, Dr. Nesil Bayraktar, Dr. Emre Vudalı, Dr. Mustafa Akansoy, Dr. Emine Kamiloğlu, Dr. Yağmur Aldağ, Dr. Hatice C. Caglayan, Dr. Fatma Canbay and Dr. Derlen O. Rus - the treating physicians who contributed to the baseline characteristics data on study participants, and finally, Dr. Ali Pilli, Dr. Ali Caygur, Dr. Sonuc Buyuk, Dr. Cigdem Caga and Dr. Figen Gulen Ince from the Health Authority in Northern Cyprus for supporting this study.

**T.K.** was supported by the Novo Nordisk Foundation Center for Protein Research (grant NNF14CC0001).

## Data Availability Statement

The datasets generated for this study are available upon request to the corresponding author.

## Ethics Statement

The study has been reviewed and approved by the International Cyprus University Ethics Committee. The COVID-19 cases i.e. the participants provided their written informed consent to participate in this study.

## Author Contributions

Elcin Yoldascan, Burc Barin, Fatma Savaskan planned the study and its implementation. Burc Barin, Huseyin Cakal, Elcin Yoldascan and Fatma Savaskan designed the psychological survey.

Fatma Savaskan coordinated the fieldwork for collection of blood samples, administration of psychological surveys and compilation of survey data.

Goncagul Ozbalikci coordinated processing of blood samples, running of immunoassays and compilation of the assay data.

Burc Barin and Huseyin Cakal performed the statistical analyses.

Burc Barin wrote the first draft.

Tugce Karaderi and Huseyin Cakal conducted critical review and editing for the major revisions.

Burc Barin, Tugce Karaderi, Huseyin Cakal conceptualized, revised and finalized the article.

All authors have reviewed the article, provided feedback and approved the article for publication.

